# Outcomes of the 2019 Novel Coronavirus in patients with or without a history of cancer - a multi-centre North London experience

**DOI:** 10.1101/2020.04.16.20061127

**Authors:** Nalinie Joharatnam-Hogan, Daniel Hochhauser, Kai-Keen Shiu, Hannah Rush, Valerie Crolley, Emma Butcher, Anand Sharma, Aun Muhammad, Muhammad Anwar, Nikhil Vasdev, Ganna Kantser, Aramita Saha, Fharat Raja, John Bridgewater, Khurum Khan

## Abstract

**Background:** Four months after the first known case of the 2019 novel coronavirus disease (COVID-19), on the 11^th^ March 2020, the WHO declared the outbreak a pandemic and acknowledged the potential to overwhelm national healthcare systems. The high prevalence and associated healthcare, social and economic challenges of COVID-19 suggest this pandemic is likely to have a major impact on cancer management, and has been shown to potentially have worse outcomes in this cohort of vulnerable patients (1). This study aims to compare the outcomes of reverse transcriptase polymerase chain reaction (RT-PCR) confirmed COVID-19 positive disease in patients with or without a history of cancer.

**Method:** We retrospectively collected clinical, pathological and radiological characteristics and outcomes of COVID-19 RT-PCR positive cancer patients treated consecutively in four different North London hospitals (**cohort A**). Outcomes recorded included morbidity, mortality and length of hospital stay. All clinically relevant outcomes were then compared to consecutively admitted COVID-19 positive patients, without a history of cancer (**cohort B**), treated at the primary centre during the same time period (12^th^ March-7^th^ April 2020).

**Results:** A total of 52 electronic patient records during the study time period were reviewed. Cohort A (median age 76 years, 56% males) and cohort B (median age 58 years, 62% male) comprised of 26 patients each. With the exclusion of cancer, both had a median of 2 comorbidities. Within cohort A, the most frequent underlying cancer was colorectal (5/26) and prostate cancer (5/26), and 77% of patients in Cohort A had received previous anti-cancer therapy. The most common presenting symptoms were cough and pyrexia in both cohorts. Frequent laboratory findings included lymphopenia, anaemia and elevated CRP in both cohorts, whilst hypokalaemia, hypoalbuminaemia and hypoproteinaemia was predominantly seen amongst patients with cancer. Median duration of admission was 7 days in both cohorts. The mortality rate was the same in both cohorts (23%), with median age of mortality of 80 years. Of cancer patients who died, all were advanced stage, had been treated with palliative intent and had received anti-cancer therapy within 13 days of admission.

**Conclusion:** Old age, late stage of cancer diagnosis and multiple co-morbidities adversely influence the outcome of patients with COVID-19 positive patients. Whilst extra caution is warranted in the administration of anti-cancer therapies pertaining to the risk of immune-suppression, this data does not demonstrate a higher risk to cancer patients compared to their non-cancer counterparts.

## INTRODUCTION

The 2019 novel coronavirus disease (COVID-19) was first detected as a case of pneumonia of unknown cause in late 2019 in Wuhan, the capital city of Hubei province in China (2). On March 11^th^ 2020, the World Health Organisation (WHO) declared COVID-19 a pandemic, due to the global spread of this new disease and the significant risk for further transmission (3). As of the 10^th^ April 2020, there were 73,758 confirmed cases in the UK, with 8,958 deaths (4).

The pathogen causing COVID-19, the severe acute respiratory syndrome coronavirus (SARS-CoV-2), is an enveloped RNA virus belonging to the family of Coronaviridae and is one of seven species of coronavirus known to infect humans. Four strains typically cause common cold symptoms and two, Severe Acute Respiratory Syndrome coronovirus (SARS-CoV) and Middle East Respiratory Syndrome coronavirus (MERS-CoV), can cause fatal respiratory diseases [Figure 1] (5).

**Figure 1:**
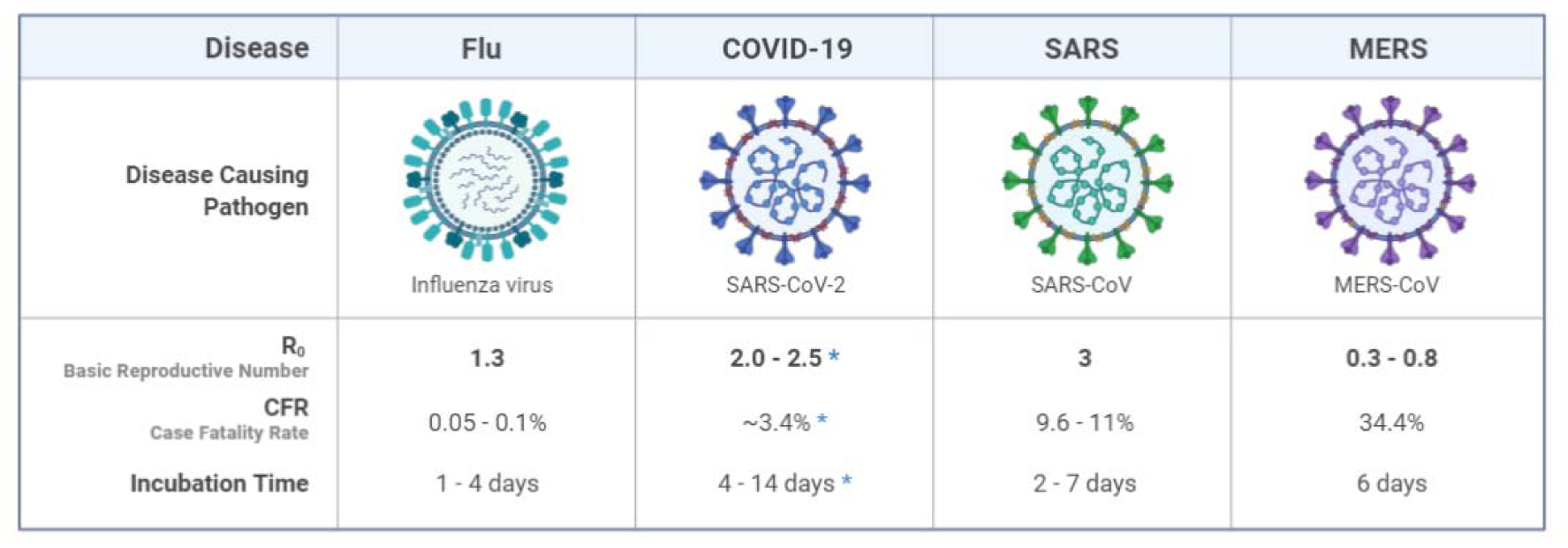
Comparison of Respiratory Viruses (6, 7) *Created with BioRender. Covid-19 data as of March 2020*

The clinical spectrum of COVID-19 ranges from asymptomatic infection to severe respiratory failure and death (8). A retrospective case study of 191 adult inpatients in Wuhan found an increased odds of death with older age (odds ratio (OR) 1.10, 95% confidence interval (CI) 1.03-1.17), as well as in those with coronary heart disease and diabetes (8). Individuals with serious chronic medical conditions, including cardiovascular disease, diabetes, lung and kidney disease are deemed vulnerable and were considered to be at higher risk of critical illness (9), including multiple organ failure and acute respiratory distress syndrome (ARDS) (10). In a prospective nationwide analysis of 1590 patients with COVID-19 in China, 18 (1%) had a history of cancer, which is higher than the prevalence of cancer in the general Chinese population. These patients were more likely to have a severe event (39% versus 8% in patients without cancer, p=0.0003), defined as the requirement of invasive ventilation or death (11). Of the cancer patients with previous history available, the majority were undergoing routine follow-up after primary resection (12 out of 16), with the rest having received chemotherapy or surgery within the past month (4 out of 16) (11). Although this is one of the larger analyses of COVID-19 in cancer patients thus far, it remains a small sample size of the total cases from China, with significant heterogeneity between diseases (12). The majority of patients were cancer survivors thus it is unclear whether these outcomes can be generalised to our oncology population, and outcome is likely to be confounded by a higher median age than those without cancer (13). A similar retrospective case study of 28 patients with a history of cancer from 3 hospitals in Wuhan found that lung cancer was the most prevalent tumour type amongst COVID positive patients (25%). Patients who had received their last anti-cancer treatment within 14 days had a significantly increased risk of developing severe events (HR=4.079, 95%CI 1.086-15.322, P=0.037) (14), highlighting the importance of carefully reviewing the necessity and priority of anticancer therapy in these high risk patients during the COVID-19 pandemic.

One of the biggest challenges in general with COVID-19 is its long incubation period [Figure 2] and potential spread through asymptomatic patients. In cancer patients, there is likely to be considerable difficulty in assessing patients for suitability for chemotherapy, as oncologists often rely only on symptoms and haematological/biochemical parameters, as well as performance status. Patients may be in the asymptomatic phase when initially assessed in clinic, and the virus peak may coincide with the myelosuppressive nadir of chemotherapy [Figure 2].

**Figure 2:**
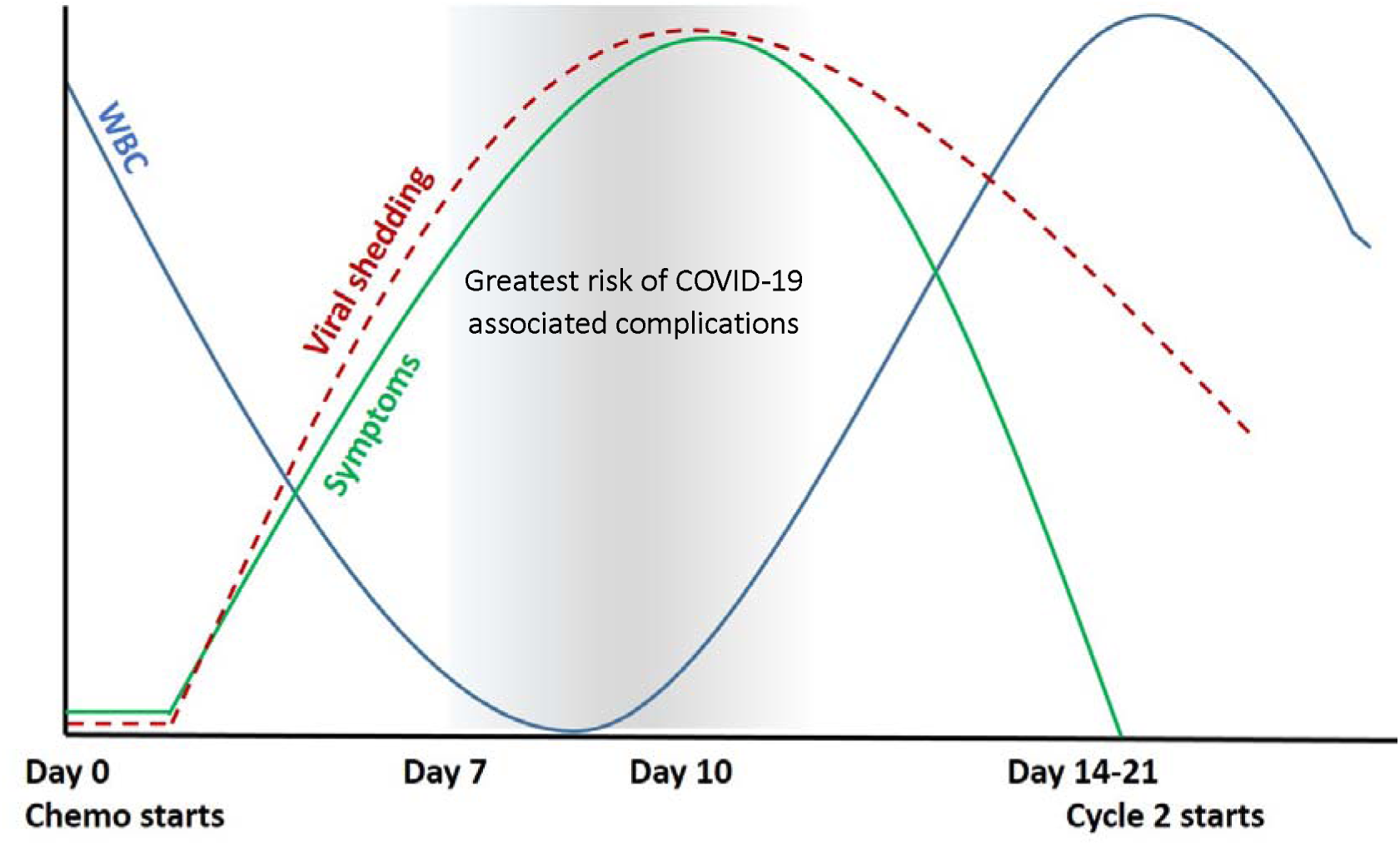
Proposed association between chemotherapy nadir and COVID-19 symptoms/viral shedding (8)

Another significant challenge for these patients is the consideration of re-starting chemotherapy in COVID-19 positive patients. It has been suggested in a case report that recurrence in patients with previously positive COVID-19 may occur, or that patients may remain persistently positive despite resolution of symptoms (15). In the case report, following initial inpatient treatment for symptomatic COVID-19 positive disease and two subsequent negative results, the patient had a recurrent positive swab result despite remaining asymptomatic. It is not yet fully understood if patients develop immunity following exposure to COVID-19, much like other coronaviruses, or if the virus can remain latent, potentially putting immunocompromised patients at a greater risk of recurrence.

Here we present a study of 26 patients with cancer alongside 26 patients without a history of cancer, admitted with COVID-19 in four North London Hospitals, to expand on the data in this setting, and to compare the differences in clinical characteristics and outcomes between these groups of patients.

## METHOD

### Patient Population

The study data includes patients admitted and diagnosed with COVID-19 between 12^th^ March and 7^th^ April 2020 at four London hospitals. We defined a confirmed diagnosis of COVID-19 based on RT-PCR from respiratory tract swabs. Cohort A comprised 26 patients with a history of cancer (both solid organ and haematological). Clinically relevant outcomes were then compared with 26 SARS-CoV-2 RT-PCR positive patients without a history of cancer consecutively admitted at the primary centre during the same time period (cohort B).

### Data Collection

Patient records were reviewed using the hospital electronic medical records (EMR) for clinical and radiological characteristics, including age, gender, comorbidities, presenting haematological and radiological characteristics and the COVID-19 management of these patients. Data specific for cohort A included site of primary tumour, intent of treatment, treatment modality and interval between last treatment and admission with COVID-19. Investigators from the different hospitals provided data of consecutive COVID-19 positive patients for cohort B at their sites and were blinded to the data from other hospitals. Severe disease was defined as a composite of intensive care admission (ITU), ventilation or death. Approval of the study was obtained from the Institutional Research and Governance team at North Middlesex University Hospital.

### Statistics

Patient demographics and clinical characteristics were explored descriptively using STATA v15.1 (StataCorp. 2017). Categorisation of numerical variables were undertaken based on consideration of the standard reference values (normal range vs low/elevated) or according to median values. Given the small number in the sample, and limited number of severe events in each cohort, it was not appropriate to examine prognostic factors and characteristics of those that died using statistical tests. Instead, frequencies and proportions (for categorical variables) or median and interquartile ranges (25^th^ percentile and 75^th^ percentile; for continuous variables) were examined for those that died by the end of the follow-up period.

## RESULTS

### Baseline characteristics of the study population

Data was collected on 28 cancer patients from four London Hospitals. 2 records were not analysed, due to duplication from readmission. Data from a further 26 patients without a history of cancer consecutively admitted with a diagnosis of COVID-19 were analysed from the primary London hospital. The median age of cohort A was 76 (interquartile range IQR 72-78), and 58 years (IQR 52-77) in cohort B. Patients were predominantly male in both cohorts, 58% (15/26) in A and 62% (16/26) in B. Excluding malignancy, comorbidity prevalence was similar between both cohorts, with a median of 2 comorbidities in each group [Table 1; and Supplementary Tables 1 and 2].

**Table 1:**
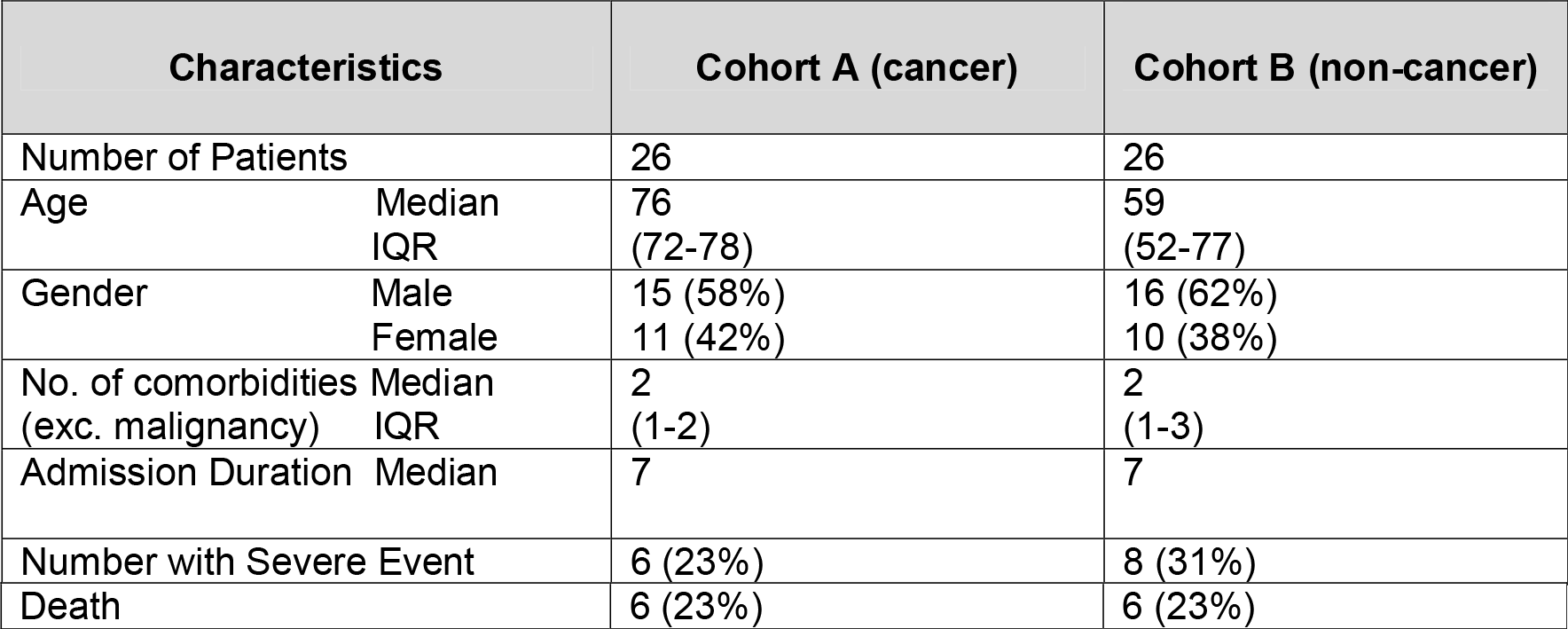
Patient Characteristics.

**Table 2:**
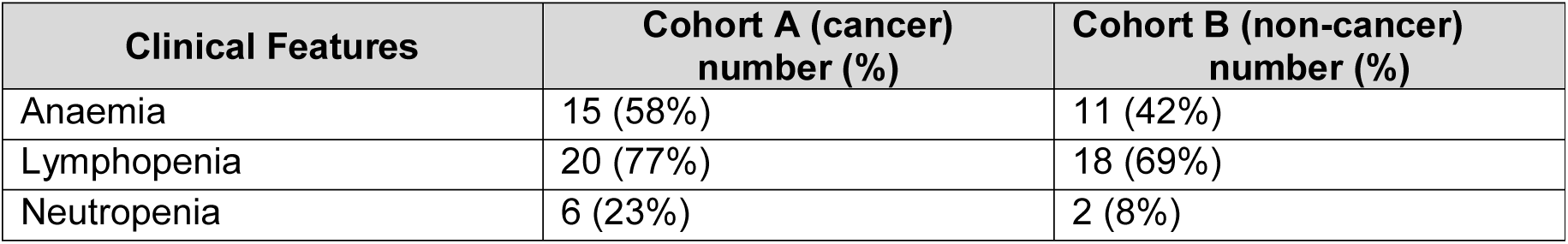
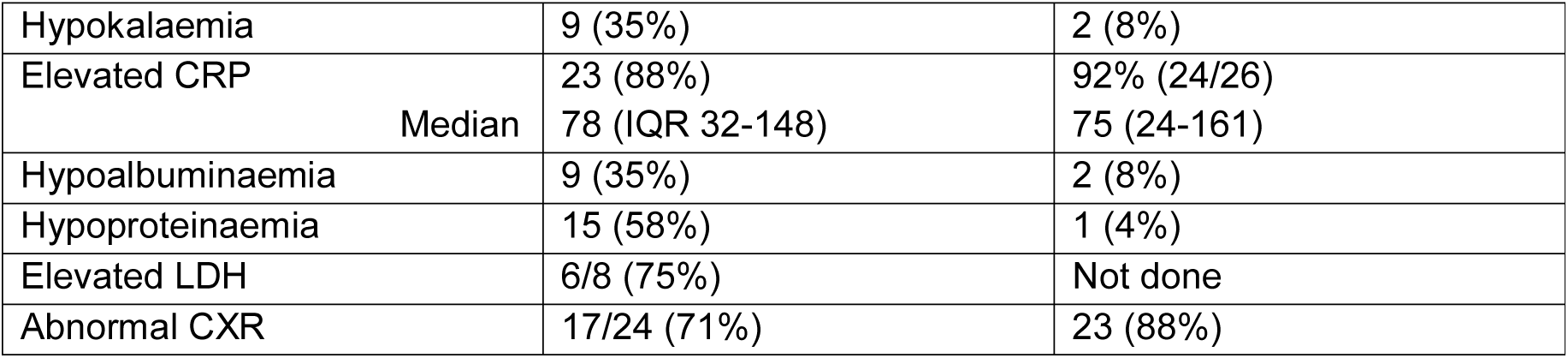
Clinical Features.

### Cancer Specific Characteristics (cohort A)

Cohort A included patients with a variety of tumour types. Colorectal and prostate cancers were the most frequent (5 of each tumour type; 19%). Most patients (77%; 20/26) had received anti-cancer therapy, 46% (12/26) chemotherapy, 27% (7/26) targeted treatment, and 1 patient had received radiotherapy. No patients had received immune checkpoint inhibitors. The majority of these patients (62%; 16/26) had received their anti-cancer therapy within 28 days of admission. 19% (5/26) of patients had not received any systemic treatment (2 of whom were planned to commence treatment in the coming weeks, 2 had been on watch and wait, and 1 had not been sufficiently fit to proceed with systemic treatment). 1 had undergone radical surgery alone. Treatment intent was palliative in 65% (17/26) of patients, all of whom had advanced stage disease. The remaining 9 patients had early stage cancers and were being treated with radical intent [Supplementary Table 1]. The median time from most recent anti-cancer treatment to presentation was 8.5 days.

### Clinical manifestations

Cough was the most frequent presenting symptom in both cohorts, 77% (20/26) and 81% (21/26) in groups A and B respectively, with pyrexia the next most common symptom in 65% (17/26) (A) and 62% (16/26) (B), as previously reported. 27% (7/26) (A) and 62% (16/26) (B) of patients were dyspnoeic on presentation. 5 (19%) cancer patients had concurrent symptoms of confusion on admission, compared with only 1 (4%) of the non-cancer patients.

### Pathological and radiological features

The haematological, biochemical and radiographic features of all 52 patients are summarised in Table 2, with full details in Supplementary Tables 3 and 4. 6 (23%) of the cancer patients were neutropenic on admission, 5 (19%) of whom had received recent anti-cancer therapy, and 2 (8%) of non-cancer patients. 77% (20/26) (A) and 69% (18/26) (B) were lymphopenic, and 58% (15/26) (A) and 42% (11/26) (B) were anaemic. Most patients in both cohorts had an abnormal C-reactive protein (CRP) with a median value of 78mg/L (A) and 75mg/L (B) (12% and 8% of cohorts A and B respectively had a normal CRP). As per previous reports of cancer patients with COVID-19, hypokalaemia was a presenting feature in 35% (9/26), hypoalbuminaemia (35%; 9/26) and hypoproteinaemia in 58% (15/26), however these were less frequent in non-cancer patients, 8% (2/26), 8% (2/26) and 4% (1/26) respectively. Only 8 patients in cohort A had a lactate dehydrogenase (LDH) tested on admission, 75% (6/8) of these were raised, ranging from 275-1800U/L.

Chest radiographs were performed in 24/26 cancer patients and all of those without cancer. These were normal at presentation in 29% (7/24) in cohort A and 12% (3/26) of cohort B. Bilateral changes were present on 53% (13/24) of abnormal chest radiographs in cohort A, and 69% (18/26) in cohort B. 38% (10/26) patients in cohort A went on to receive a CT chest, of which 7 (27%) demonstrated bilateral basal or peripheral changes (1 patient had a normal chest CT). Only 1 patient in cohort B had a CT chest, demonstrating bilateral atelectasis.

### Outcomes of the study population and its association with relevant clinic-pathological parameters

The median hospital stay for patients admitted with COVID-19 in both cohorts was 7 days. One patient in the cancer case series developed likely ‘nosocomial’ COVID-19, having been admitted for an unrelated cause of bowel obstruction, although had not been swabbed for COVID-19 on admission.

All cancer patients received ward-based supportive care only. Supportive treatment generally consisted of intravenous fluids and antibiotics. None of these patients were admitted to ITU or treated with invasive ventilation. 12% (3/26) in cohort A received non-invasive ventilatory support (continuous positive airway pressure - CPAP), all of whom subsequently died. In total 23% (6/26) patients died in cohort A, with no other cases meeting the predefined markers of severity.

Within the non-cancer patients, 19% (5/26) of patients were admitted to ITU, and 23% (6/26) patients died. Overall 31% (8/26) met the predefined marker of severity (ventilation, ITU admission or death) [Table 1].

The median age in those who died was 80 years in both cohorts. Of those who died most cancer patients were male (83%) and 50% male in cohort B. Although cohort B had a higher rate of non-cancer comorbidities in those who died, with inclusion of malignancy both groups had similar rates, with the median number of comorbidities in those who died 3 in both cohorts.

### Cancer Specific Outcomes

Of the 6 patients with cancer who died, all had received anti-cancer therapy within 13 days of admission. 50% (3/6) had received chemotherapy, and 50% (3/6) had received targeted treatment. All 6 patients deteriorated and died within 14 days of admission. 4 had a history of prostate cancer, 1 pancreatic and 1 extensive small cell lung cancer. The median time interval between anti-cancer therapy and admission was 1 day (IQR 1-8) in those who died, compared to 12.5 (IQR 7-29) days in those who survived. All patients who died were being treated with palliative intent or in the non-curative setting of cancer. In this cohort, there was little difference in the median age between those who died (median 80 years, IQR 76-86) and those who survived (median 75 years, IQR 71-78). 100% (6/6) were lymphopenic, with a raised CRP on presentation. Neutropenia was present in 50% (3/6), 100% (6/6) had hypoproteinaemia and 83% (5/6) were hypokalaemic on admission. 83% (5/6) had abnormal chest radiograph findings, of which 67% (4/6) were bilateral.

## DISCUSSION

To our knowledge, this is the first report comparing the outcomes of COVID-19 positive patients with a history of cancer with their non-cancer counterparts. This study demonstrates that COVID-19 infection carries a significant risk of morbidity and mortality for all patients regardless of their history of malignancy.

The results of the oncology specific patients are comparable to published literature from a similar series of patients with cancer and COVID-19 in Wuhan, China (14). In this series of 26 cancer patients treated in 4 London hospitals, none received invasive ventilation or intensive care admission and 6 died (23%), with a comparable mortality in the non-cancer group. The cancer cohort was comprised of a predominantly older population, with a median age of 76 years (unlike the cohort published in Wuhan, in whom the median age was 65) (14). This may reflect the older age of cancer patients in the UK. The non-cancer cohort in this study were a younger group, with a median age of 58, similar to the average age of patients generally admitted to hospitals in the UK (16).

Clinical features of both cohorts of patients were consistent with previous studies, with cough and pyrexia the most frequent presenting symptoms. 5 cancer patients, compared to 1 non-cancer patient, had concurrent confusion. This may be attributed to hypoxia, but also perhaps be a reflection of the older age, and therefore higher risk of delirium, in this cohort. Typical laboratory features in both groups included anaemia, lymphopenia and a raised CRP, however patients with cancer more frequently had features of hypoalbuminaemia, hypoproteinaemia and hypokalaemia (the former two may reflect possible malnutrition associated with malignancy, advanced disease and frailty). These were features also reported by the cohort of cancer patients in Wuhan by Zhang et al (14), and therefore recognition of these features may help to identify patients at highest risk of deterioration.

The mortality in each cohort of patients was 23% (6/26), significantly higher than the current estimation of mortality in hospitalised patients (>5%) (17, 18); however both groups of patients had significant co-morbidities, of the same median number (3). A history of advanced malignancy did not appear to increase the mortality rate compared to those without cancer, acknowledging that this cohort study has a small sample size. Of the 6 cancer patients who died, all had received anti-cancer therapy, and all in the palliative setting of advanced disease. Patients were more likely to die if treatment had been administered within 14 days of admission, corroborating Zhang *et al*.’s dataset(14).

Our data show that outcomes between cancer and non-cancer patients were comparable. Whilst patients with cancer tend to have underlying immunosuppression from anti-cancer therapies and sometimes due to the underlying malignancy itself, and therefore perhaps more inclined to contract infection, cytokine storm syndrome might be the primary reason for mortality in COVID-19 positive patients more generally (19). Contrary to the general hypothesis that patients on immunosuppressive regimes such as chemotherapy are at particular risk of adverse outcomes after contracting COVID-19, immunosuppression may paradoxically work in their favour, by dampening this cytokine storm response. A recent study suggested use of immune-suppressive therapies such as steroids, intravenous immunoglobulin, selective cytokine blockade (eg, anakinra or tocilizumab) and JAK inhibition might act as potential therapeutic options (19).

Based on the data presented here, there was no excess in mortality in the cancer cohort despite many patients being actively treated with anti-cancer therapy. However all the patients in the cancer cohort who died had received systemic anti-cancer therapy (SACT) within the last 14 days. Thus cautious consideration should be made of the risk/ benefit profile of palliative SACT in the setting of the COVID-19 pandemic to minimise the potential risks of mortality in already vulnerable patients.

We acknowledge the limitations to this study, including the retrospective sample collection and small sample size. Amongst the patients with cancer, there was significant heterogeneity between tumour types, including variability in stage and other clinico-pathological factors. Patients were selected consecutively, rather than choosing matched samples between cohorts, leading to possible confounding. Patient data was obtained from 4 different London hospitals, in which treatment may vary, and this therefore may have an impact on prognostic factors. However, selecting admitted patients in a consecutive manner helped to reduce selection bias. Selection bias was further reduced by selecting data from the primary institute only in the non-cancer ‘control’ arm. Data were date matched; however not age or gender matched. Given the limited sample size with a pre-defined severe event, analysis of the effects of multiple prognostic risk factors on mortality could not be carried out, due to the likely effect of other confounding variables, and this would be a useful analysis to undertake in a larger series of patients. We acknowledge the above limitations. Given the urgent timeline of an evolving and rapidly progressing pandemic, we elected not to wait for a larger sample-size, as we prioritise the early release of data to assist global physicians and oncologists to make real-time decisions.

In the UK, the government has closed all non-essential places of work, limiting travel to key-workers only and encouraging social distancing (20). It is thought that the progression of infection in the UK is approximately two weeks behind Italy, with 139,422 confirmed cases and 17,669 deaths at the time of writing (21). Due to the likely significant burden on the NHS in the coming weeks to months, and the potential risks for cancer patients, the prioritisation of chemotherapy has been recommended to meet capacity, mitigate the risks of COVID-19, and to allow patients to have the best opportunity for cure or disease control. Over the last 3 weeks a number of recommendations have been published to guide the oncological management of patients with COVID-19, and suggestions for how to prioritise treatment based on resources, but also how best to identify those most vulnerable. A recent publication in the Lancet Oncology succinctly summarises published guidelines for cancer during the COVID-19 pandemic (22). Both cohorts in this study had a number of underlying comorbidities, which has been identified to be a significant risk factor for severity in COVID-19 (8), however this study did not find that a history of malignancy increased the severity of outcomes from COVID-19.

## CONCLUSION

COVID-19 is a significant cause for health-care, societal and economic concern globally. Previous work has demonstrated older patients, and those with significant comorbidity, are at the greatest risk of morbidity and mortality from the virus. Patients with cancer are at higher risk of immunosuppression on cancer therapy, with possible increased incidence of infection in these patients based on data from China. In our series of patients, the potential increased risks of mortality appear specific to those patients with more advanced cancer, those who were older, and those receiving recent systemic anti-cancer therapy. However, compared to patients without a history of cancer, patients with cancer were no more likely to have a severe outcome or mortality from the infection. Whilst extra caution is warranted in administration of anti-cancer therapies pertaining to risk of immune-suppression, this data does not demonstrate a higher risk to cancer patients compared to their non-cancer counterparts. However further patient data is required to better understand the appropriate management of these patients, in our potentially increasingly overwhelmed healthcare system.

## Data Availability

Datasets generated or analysed during the current study are available on request

